# The Impact of Omega-3 Fatty Acids on SM23-33 Abundance in Stool and Children with Obesity: A Mendelian Randomization Analysis

**DOI:** 10.1101/2024.07.01.24309764

**Authors:** Min Zhang

## Abstract

**Background:** Children with obesity may be associated with gut microbiota and omega-3 fatty acids. However, the understanding of children with obesity, gut microbiota, and omega-3 fatty acids remains unclear.

**Objectives:** This study aimed to examine the relationships between omega-3 fatty acids, SM23-33 abundance in stool, and children with obesity.

**Methods:** We conducted LDSC to assess the genetic correlation between SM23-33 abundance in stool, Omega-3 fatty acids, and children with obesity. Subsequently, bidirectional MR analyses were performed to investigate the causal connections between SM23-33 abundance in stool and children with obesity, while a two-step MR analysis was employed to identify any potential mediation by Omega-3 fatty acids in this relationship. All statistical analyses were carried out using R software, and the STROBE-MR checklist was followed for reporting MR studies.

**Results:** There was no significant genetic correlation among SM23-33 abundance in stool, Omega-3 fatty acids, and children with obesity (*rg_p*>0.05). MR analysis identified SM23-33 abundance in stool causally associated with children with obesity (*OR*=0.747, 95%*CI*: 0.584-0.957, *P*=0.021). Furthermore, there was no strong evidence that genetically predicted children with obesity affected SM23-33 abundance in stool. Further, two-step MR analysis found the associations between SM23-33 abundance in stool and children with obesity were mediated by Docosahexaenoic Acid (DHA) of Omega-3 fatty acids with proportions of 3.56% (95%*CI*: 3.43%, 3.69.0%).

**Conclusions:** The present study provides evidence supporting the causal relationships between SM23-33 Abundance in Stool and Children with Obesity, with a potential effect mediated by Docosahexaenoic Acid (DHA).

## 1 INTRODUCTION

Obesity is a chronic complex disease defined by excessive fat deposits that can impair health. Obesity can lead to an increased risk of type 2 diabetes and heart disease; it can affect bone health and reproduction, and it increases the risk of certain cancers.^1^ Obesity influences the quality of living, such as sleeping or moving.^2^ According to the latest data from the World Health Organization, in 2022, 1 in 8 individuals worldwide had obesity. Since 1990, adult obesity has more than doubled, and adolescent obesity has quadrupled. By 2022, over 390 million children and adolescents aged 5-19 years had overweight, including 160 million who had obesity. These statistics highlight the severity of the obesity epidemic and the need for effective strategies to address this global health issue.

The gut microbiome plays a crucial role in the development and progression of obesity.^4^ The gut microbiome has been increasingly recognized as a crucial factor in the development and progression of children with obesity. Research has shown that an altered gut microbiome is associated with children with obesity, characterized by a reduced diversity of beneficial bacteria and an increased abundance of opportunistic pathogens. Studies have consistently demonstrated that children with obesity have a distinct gut microbiome profile compared to lean children. For example, the Firmicutes phylum is enriched in children with obesity, while the Bacteroidetes phylum is enriched in lean children.^5^ Additionally, the gut microbiome of children with obesity has been shown to be less diverse and more prone to inflammation.^6^ The gut microbiome plays a crucial role in regulating energy harvest and storage, and alterations in the gut microbiome can lead to changes in metabolic pathways, contributing to the development of obesity in children.^7^ Furthermore, the gut microbiome has been shown to influence the regulation of appetite and satiety hormones, such as ghrelin and leptin, which can contribute to overeating and weight gain in children.^8^ Modulating the gut microbiome through dietary interventions, such as prebiotics or probiotics, has been shown to be an effective strategy for weight management in children.^9^

Despite the established link between the gut microbiome and children with obesity, there is still much to be learned about the specific contributions of different microbial families to this relationship. The SM23-33 order, comprising strictly fermentative Phycisphaerae bacteria, is one such family that warrants further investigation. The SM23-33 order currently comprises two families, including SM23-33 and FEN-1343. They were found in metagenomic datasets obtained from estuary sediments, sulphur-rich hydrothermal sediments,^10^ and anaerobic digesters,^11^ but literature descriptions of this order are scarce. Recent research indicates that the functional capacity of the SM23-33 family of bacteria may be more complex than currently understood. These bacteria not only thrive in environments devoid of electron acceptors but also have the ability to degrade complex carbon substrates. One notable finding is that certain members of this family may possess the ability to perform anaerobic sulfite reduction, highlighting their metabolic diversity and ecological adaptability. ^12^ Although there is currently no direct evidence linking SM23-33 bacteria to children with obesity, their diverse metabolic capabilities and potential roles in complex carbon metabolism suggest that further research is warranted. Such studies could provide deeper insights into their potential impacts on human health and microbial community structures, possibly uncovering connections to complex diseases, including children with obesity.

While the potential links between SM23-33 bacteria and children with obesity warrant further investigation, other research has focused on the role of nutrition in preventing and managing this disease, including the potential benefits of omega-3 fatty acids. Omega-3 fatty acids, particularly Alpha-linolenic Acid (ALA), Docosahexaenoic Acid (DHA), Docosapentaenoic Acid (DPA), and Eicosapentaenoic Acid (EPA), may play a beneficial role in combating children with obesity. These fatty acids have demonstrated the ability to modulate inflammation, enhance insulin sensitivity, and improve cardiovascular health. ^13 - 15^ Additionally, Omega-3 fatty acids may influence appetite regulation and satiety, leading to reduced energy intake and better weight management.^16^ Two systematic reviews and meta-analyses conducted in 2019 found that omega-3 fatty acid supplementation significantly reduced body mass index (BMI) and waist circumference in children and adolescents. ^17^ Another 2020 study reported improvements in insulin sensitivity and reductions in inflammatory markers among children with obesity receiving Omega-3 supplements.^18^ However, not all findings are consistent. Some studies have yielded inconclusive or even contradictory results. For example, A 2019 review of 22 clinical trials found no significant effect of omega-3 fatty acid supplementation on children’s BMI or body fat percentage.^19^ A 2020 systematic review and meta-analysis confirmed these findings, reporting no substantial impact from omega-3 supplements.^20^ Additionally, a 2018 study observed an increase in BMI and body fat percentage in children with obesity following Omega-3 supplementation.^21^ These mixed results highlight the necessity for further research to clarify the relationships between omega-3 fatty acids, gut microbiota, and children with obesity.

Mendelian randomization (MR) analysis is a robust epidemiological research strategy grounded in the principles of Mendelian inheritance. This technique allows for the estimation of causal relationships by using genetic variants as instrumental variables, thereby mitigating issues related to confounding variables, measurement errors, and reverse causation.^22^ Based on recent studies, we hypothesize that there may be a causal association between SM23-33 abundance in stool and children with obesity, with omega-3 fatty acids acting as mediators. Accordingly, we applied mediation MR analysis to investigate this relationship, using genetic data from large-scale genome-wide association studies (GWAS). Our aim is to elucidate the causal pathways involved, providing a clearer understanding of how SM23-33 influences children with obesity through its impact on omega-3 profiles.

## 2 MATERIALS AND METHODS

### 2.1 Study design

In this study, we employed a range of Mendelian Randomization (MR) methodologies, including genetic correlation analysis (LDSC), two-sample MR (TSMR), bidirectional MR (BDMR), multivariable MR (MVMR), and two-step MR (2SMR), to investigate the causal relationships between SM23-33 abundance in stool, omega-3 fatty acids, and children with obesity. Our study design illustrated is based on three key assumptions essential for the causal interpretation of MR estimates.^23^ We ensured the reliability of our findings by using genetic variations or single nucleotide polymorphisms (SNPs) as instrumental variables that meet three crucial criteria: (i) the genetic IVs are strongly associated with exposure, (ii) the genetic IVs are not associated with confounders linked to the selected exposure and outcome, and (iii) the genetic IVs influence the outcome only through the exposure. By employing multiple MR methods, we minimized bias and obtained reliable estimates of modifiable exposures and their relationships with targeted outcomes. Additionally, we completed the STROBE-MR checklist to ensure the integrity of this observational MR study; see supplementary material (Data S1, Table S1) for details. Figure 1 provides a clear and transparent overview of our study design.

**Figure 1.**
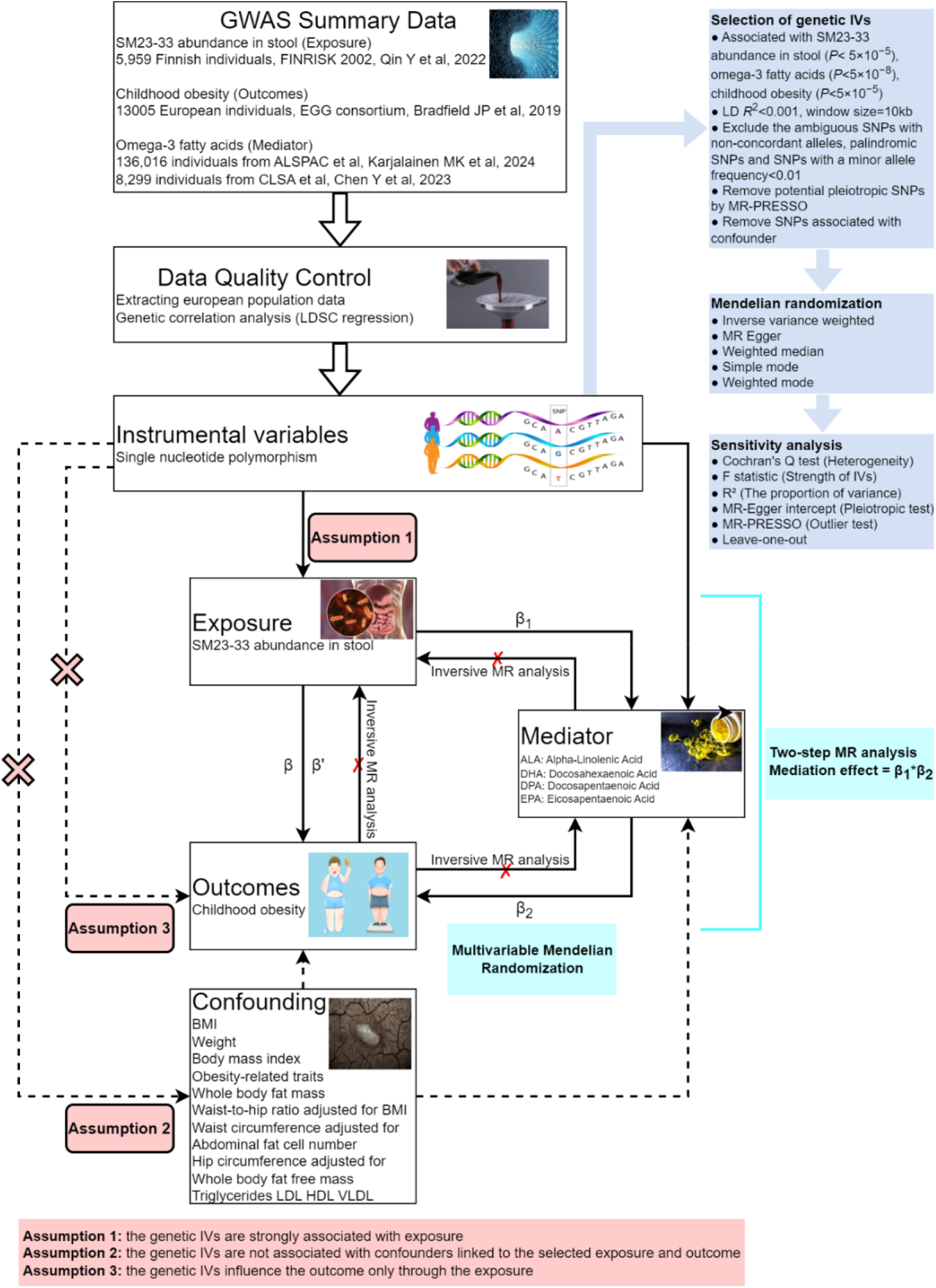
Overall study design plan. GWAS, genome-wide association studies; SNPs: single-nucleotide polymorphisms; β: the total causal effects of exposure on outcome was calculated using UVMR method; *β*_1_: the causal effects of exposure on mediators were calculated using UVMR method. *β*_2_: the causal effects of mediators on outcome were calculated using MVMR method. *β*’: the direct causal effects of exposure on outcome were calculated using the formula *β’=β-(β*_*1*_*×β*_*2*_).

### 2.2 Data sources

#### 2.2.1 Source of SM23-33 Abundance in Stool

SM23-33 abundance in Stool association studies (GWAS) data utilized in this research was obtained from a study of genetic characteristics of gut microbiota; see supplementary materials for details (Data S2, Table S3). The original GWAS was conducted on 2,801 microbial taxa and 7,967,866 human genetic variants from 5,959 individuals enrolled in the FR02 cohort. GWAS summary data for SM23-33 abundance in Stool was downloaded from GWAS Catalog (https://www.ebi.ac.uk/gwas), and the GWAS Catalog accession number is GCST90032559. More detailed information about the GWAS data can be obtained from their study.^24^

#### 2.2.2 Source of Omega-3 fatty acids

The GWAS data for DHA was sourced from the GWAS Catalog originally generated by Karjalainen MK et al.,^25^ with the GWAS Catalog accession number GCST90301955. This comprehensive study identified genetic associations for 233 circulating metabolic traits across 33 cohorts, encompassing a total of 136,016 participants, predominantly of European ancestry, with a small proportion from Asia (11.60%,15775/136016). Other omega-3 fatty acids, including ALA, DPA and EPA were sourced from the GWAS catalog originally generated by Chen Y et al.,^26^ with the GWAS Catalog accession numbers GCST90199748, GCST90199713, and GCST90200349 respectively. This study was based on the Canadian Longitudinal Study on Aging (CLSA) cohort; researchers analyzed data on 1,091 blood metabolites and 309 metabolite ratios by examining a total of 8,299 participants and approximately 15.4 million SNPs. Detailed information for the Omega-3 fatty acids were provided in the supplementary materials (Data S2, Table S3). More detailed information about the GWAS data can be obtained from their study too^2526^.

#### 2.2.3 Source of Children with Obesity

Data on children with obesity has been contributed by the EGG consortium and has been downloaded from www.egg-consortium.org. ^27^ To identify additional genetic variants associated with children with obesity, the researchers conducted a trans-ancestral meta-analysis of 30 studies, encompassing up to 13,005 cases and 15,599 controls from European, African, North/South American, and East Asian ancestries. Following screening, European-specific GWAS data were extracted for analysis. See supplementary materials (Data S2, Table S3).

### 2.3 Selection of IVs and data harmonization

To adhere to the stringent criteria based on the three principal MR assumptions and to mitigate horizontal pleiotropy, only independent genome-wide significant single nucleotide polymorphisms (SNPs) were employed as instrumental variables (IVs) for the exposure. T he IVs must be closely related to the exposure (SM23-33 abundance in stool, omega-3 fa tty acids, children with obesity), SNPs significantly associated with the occurrence were selected at the whole-genome level (*P*<5×10^−8^, *r*^2^<0.001, window size=10kb), If there are t oo few SNPs Included, the inclusion criteria can be changed to (*P*<5×10^−5^ or *P*<1×10^−5^, *r* ^2^<0.001, window size=10kb). Additionally, we calculated the *F* statistics of the IVs to assess the extent of weak instrument bias. To reduce the bias caused by weak working variables, the working variables with *F* > 10 are retained, and the SNPs with *F*>100 are retained for data sets with more SNPs. The formula for calculating the *F* value and *R*^2^ is as follows. *R*^2^=2×*β*_*exp*_^2^×*eaf*_*exp*_(1−*eaf*_*exp*_)/[2×*β*_*exp*_^2^×*eaf*_*exp*_×(1−*eaf*_*exp*_)+2×*n*_*exp*_×eaf_*exp*_×(1-eaf_*exp*_)×*se*_*exp*_ ^2^], *F*=*R*^2^×[*n*_*exp*_−2/(1−*R*^2^)].

In order to avoid multicollinearity and confounding effects, we used the LDlink tool (https://ldlink.nih.gov/?tab=ldtrait) to perform linkage disequilibrium (LD) pruning on the SNPs. Utilizing the LDtrait module, we selected the European population based on the CRCH38 genome build with *r*^2^=0.001, window size 10,000 kb. This approach ensured that one representative SNP was retained per window, thereby reducing confounding factors introduced by LD. Additionally, SNPs associated with phenotypes such as BMI, weight, body mass index, obesity-related traits, whole body fat mass, waist-to-hip ratio adjusted for BMI, waist circumference adjusted for BMI, abdominal fat cell number, hip circumference adjusted for BMI, whole body fat-free mass, triglycerides, LDL, HDL, and VLDL were removed to eliminate potential confounding. By excluding SNPs linked to these phenotypes, we aimed to reduce confounding effects and enhance the robustness of our findings. Finally, after data filtering, horizontal pleiotropy analysis was conducted. If there is horizontal pleiotropy, the MR-PRESSO test is performed. The outliers obtained from the test were removed before proceeding with further analysis.

### 2.4 Causal effects of SM23-33 abundance in stool on children with obesity

We performed bidirectional MR analyses to investigate the causal relationship between SM23-33 abundance in stool and children with obesity, estimating the total effect (β) of this relationship. We used the inverse variance weighted (IVW) method to estimate effects, reporting β±*SE* for continuous outcomes and *OR* (95%*CI*) for binary outcomes. In brief, the IVW method meta-analyzed SNP-specific Wald estimates by dividing the SNP-outcome association by the SNP-exposure association, using random effects to derive the final causal effect estimate. Additionally, we used MR-Egger and weighted median methods as complementary approaches to IVW, providing a more comprehensive understanding of the causal relationship.

### 2.5 Mediation analyses link “SM23-33 abundance in stool–Omega-3 fatty acids– children with obesity”

We further performed a mediation analysis using a two-step MR study to explore whether Omega-3 fatty acids mediate the causal SM23-33 abundance in stool to children with obesity.^28^ The specific approach is: (step 1) a univariable MR(UVMR) model was carried out to estimate the effect of the exposure on the mediator, and (step 2) a second model estimating the effect of each mediator on the outcome was carried out using MVMR. Both the genetic variants for the mediator and the exposure were included in the first and second-stage regressions in MVMR. Using MVMR ensures that the mediator’s effect on the outcome is independent of the exposure. Additionally, this method provides an estimate of the direct effect of the exposure on the outcome. The two regression estimates from the second stage regression are multiplied together to estimate the indirect effect^**Error! Bookmark not defined**.^.

With respect to our research, the overall effect can be decomposed into a direct effect (without mediators) and an indirect effect (through mediators). The total effect of SM23-33 abundance in stool on children with obesity was decomposed: direct effects of SM23-33 abundance in stool on children with obesity (overall effect) and indirect effects mediated by SM23-33 abundance in stool through the mediator (mediation effect). The mediation effect was calculated through β_1_×β_2_ :(i) the causal effect of the mediator (Omega-3 fatty acids) on the outcome (children with obesity) adjusted for exposure-induced confounding (β_2_) and (ii) the causal effect of the exposure (SM23-33 abundance in stool) on the mediator (β_1_). We calculated the percentage mediated by the mediating effect by dividing the indirect effect by the total effect. Meanwhile, 95%CI was calculated using the Delta method.

### 2.6 Sensitivity analysis

The directional association between each identified SNP and both the exposure and outcome variables was assessed using MR Steiger filtering. This method measures the degree to which the variation in exposure and outcomes can be attributed to instrumental SNPs and determines if the variability in outcomes is less than that in exposure. Horizontal pleiotropy was further investigated via the MR-Egger approach, which utilizes weighted linear regression with an unconstrained intercept. This intercept acts as an indicator of the average pleiotropic effect across genetic variations, reflecting the typical direct influence of a variant on the outcome variable. If the intercept significantly deviated from zero (MR-Egger intercept *P*<0.05), it indicated the presence of horizontal pleiotropy. Moreover, Cochrane’s Q-test was used to assess heterogeneity, with lower p-values suggesting increased heterogeneity and a higher probability of directional pleiotropy. Leave-one-out analyses were also performed to identify potential SNP outliers.

### 2.7 Statistical analysis

We first used the results calculated by the IVW method as the final main result, and a significance threshold of *P*<0.05 was applied to the MR analysis, where P-values below this threshold were considered statistically significant. All statistical analyses and data visualizations were performed using R software (R Foundation, Vienna, Austria), with the TwoSampleMR (https://github.com/MRCIEU/TwoSampleMR) package for UVMR and MVMR analysis, the GenomicSEM (Yen-Tsung Huang, Patrick J. Smith, USA) package for LDSC analysis, and the PNG (Boutell, Netherlands) package for data visualization.

We utilized LDSC to estimate the genetic correlation (rg) among SM23-33 abundance in stool, Omega-3 fatty acids, and children with obesity. LDSC assesses the relationship between test statistics and linkage disequilibrium, allowing us to quantify the contribution of inflation from a genuine polygenic signal or bias. This approach enables the evaluation of genetic correlation using GWAS summary statistics and remains unbiased by sample overlap. We computed the z-scores for each variant of Trait 1 multiplied by those of Trait 2, and the genetic covariance was derived by regressing this product against the LD score. The resultant genetic covariance, normalized by SNP-heritability, indicates the genetic correlation. Statistical significance was set at *P*<0.05.

### 2.8 Ethical statement

This research utilized publicly accessible data from GWAS studies. Each individual study included in the GWAS was approved by the respective Institutional Review Board, and participants or their authorized representatives provided informed consent.

## 3 RESULTS

### 3.1 Selection of IVs

The remaining 73 SNPs of SM23-33 abundance in stool and 120 SNPs of Omega-3 fatty acids that meet assumptions 1, 2, and 3 were screened. Meanwhile, we excluded SNPs associated with obesity, namely rs147608546, rs1995755, rs41264899, rs75596315, rs7581869, rs7791602, rs79131883, and rs9783388, as well as SNP rs56154844 associated with omega-3 fatty acids. These exclusions were based on previous evidence suggesting these SNPs may act as confounders, potentially influencing the causal inference of SM23-33 abundance in stool, omega-3 fatty acids, and children with obesity. The *F*-statistics for all instrumental variables (IVs) were above 10, indicating no evidence of weak instrument bias. Specifically, the average *F*-value for the relationship between exposure and outcome was 18.85; for exposure and mediator, it was 18.84; and for mediator and outcome, it was 162.73. These values suggest that the instrumental variables used in the analysis are strong and reliable. See supplementary materials for details (Data S2, Table S4).

### 3.2 LDSC regression analysis

We performed LDSC regression analysis to evaluate the genetic correlation among SM23-33 abundance in stool, Omega-3 fatty acids, and children with obesity. The genetic correlation analysis results revealed no significant genetic correlation among the three traits (*rg_p*>0.05), which helps us further carry out the MR analysis, as shown in Table 1. See supplementary materials for details (Data S2, Table S2).

**Table 1.** Genetic Correlations among SM23-33 Abundance in Stool, Omega-3 Fatty <u>Acids, and Children with Obesity</u>.

| Trait1 | Trait2 | $R_g$ | $R_g\_se$ | $R_g\_p$ |
| --- | --- | --- | --- | --- |
| SM23-33 Abundance in Stool | Children with Obesity | 0.042 | 0.232 | 0.858 |
| SM23-33 Abundance in Stool | Alpha-linolenic Acid | 1.280 | 0.677 | 0.059 |
| SM23-33 Abundance in Stool | Docosahexaenoic Acid | 0.253 | 0.173 | 0.144 |
| SM23-33 Abundance in Stool | Docosapentaenoic Acid | 0.612 | 0.589 | 0.299 |
| SM23-33 Abundance in Stool | Eicosapentaenoic Acid | 0.064 | 0.631 | 0.919 |
| Alpha-linolenic Acid | Children with Obesity | 0.143 | 0.156 | 0.360 |
| Docosahexaenoic Acid | Children with Obesity | -0.025 | 0.047 | 0.598 |
| Docosapentaenoic Acid | Children with Obesity | 0.132 | 0.137 | 0.338 |
| Eicosapentaenoic Acid | Children with Obesity | 0.061 | 0.139 | 0.661 |

### 3.3 Causal association between SM23-33 abundance in stool with children with obesity

62 SNPs have been used to genetically proxy the effect of SM23-33 abundance in stool (Supplementary Table S4). SM23-33 abundance in stool was negatively correlated with the risk of children with obesity (see Figure 2). The odds ratio (*OR*) was 0.747 (95%*CI*: 0.584-0.957; *P* = 0.021) for children with obesity per 1 SD increase of SM23-33 abundance in stool using inverse-variance weighted methods (see Figure 3). That is to say, for each SD increase in SM23-33 abundance in stool, the relative odds of children with obesity were 25.3% lower. See supplementary materials for details (Data S2, Table S5).

**Figure 2.**
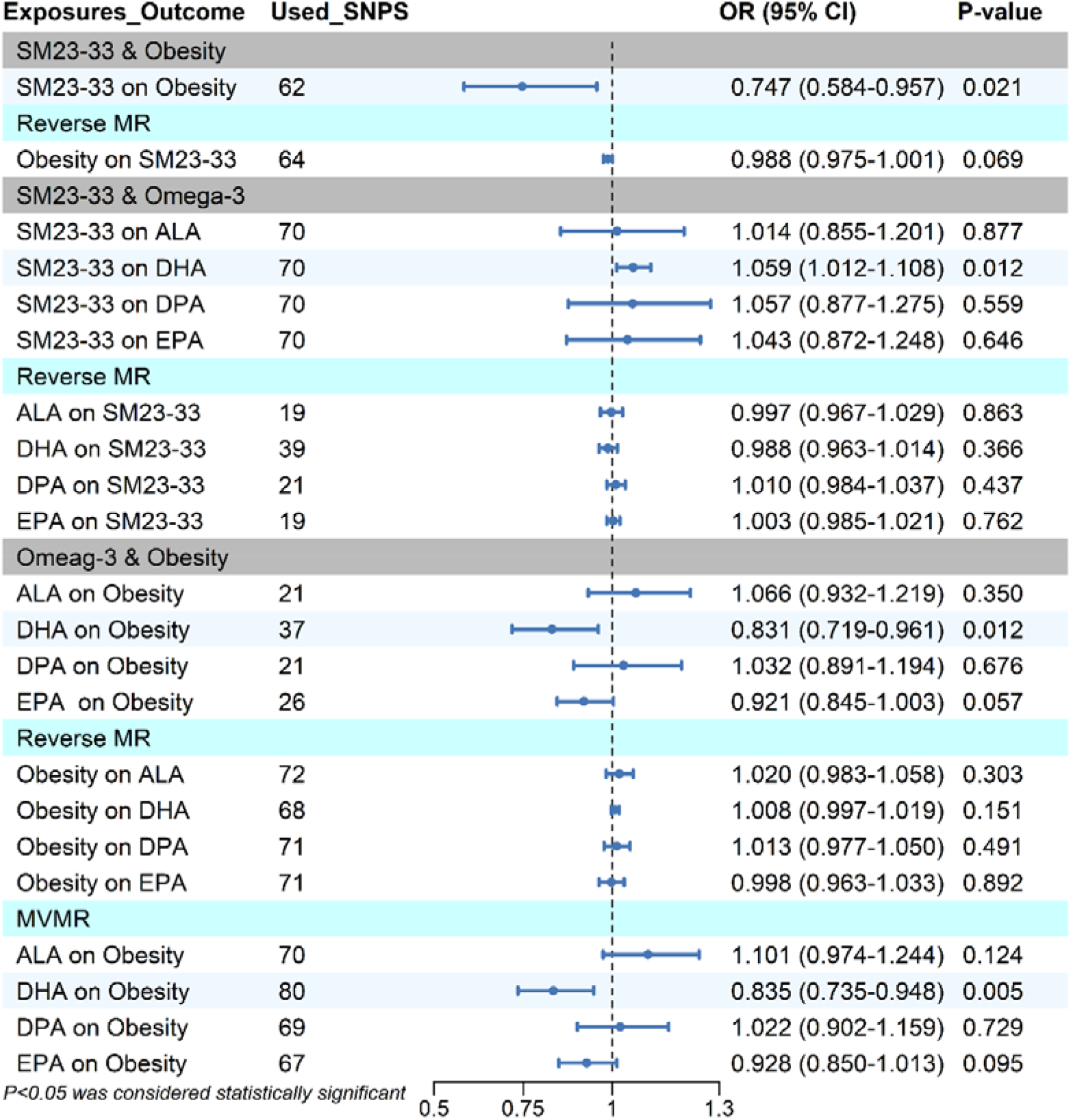
Univariable MR analysis shows causal effects of genetically proxied SM23-33 abundance in Stool, Omega-3 fatty Acids, and children with obesity using IVW methods. Abbreviations: SNPs, single-nucleotide polymorphisms; OR, odds ratio; CI, confidence interval; SM23-33, SM23-33 abundance in stool; Obesity, children with obesity; MR, Mendelian randomization; ALA, Alpha-linolenic Acid, DHA, Docosahexaenoic Acid; DPA, Docosapentaenoic Acid.

**Figure 3.**
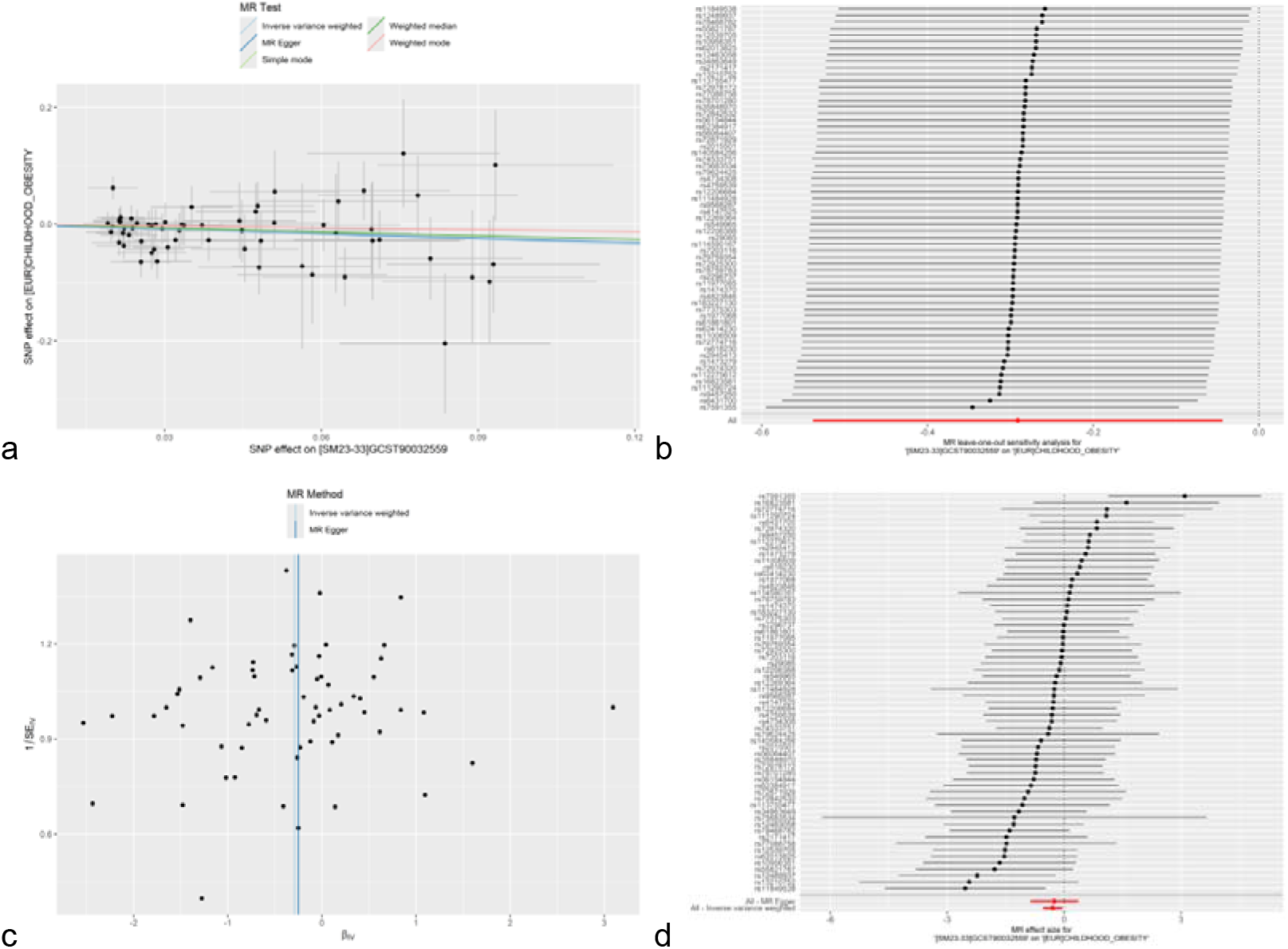
Casual effect of SM23-33 abundance in stool on childhood obesity. (a) Scatter plot illustrating the relationship between abundance in stool and childhood Obesity. The five methods utilized in this study are depicted, with lines in light blue, dark blue, light green, dark green, and red representing IVW, MR-Egger, Simple model, Weighted median, and Weight mode methods. (b) Leave-one-out analyses were conducted to ascertain whether any individual instrumental variable was exerting a disproportionate influence on the estimated causal effect. (c) The funnel plot was employed to evaluate the presence of potential heterogeneity in the observed association. (d) The forest plot was utilized to visually present the MR estimate and corresponding 95% confidence interval values for each SNP. Furthermore, the IVW and all-MR Egger results are presented at the bottom of the plot. Abbreviations: MR, Mendelian randomization; SNP, single-nucleotide polymorphism.

### 3.4 Mediation MR analyses of Omega-3 fatty acids

Genetically proxied SM23-33 abundance in stool was significantly associated with children with obesity [*OR*=0.747, (95%*CI*, 0.584, 0.957), *P*=0.021]. To investigate the potential mechanisms through which SM23-33 abundance in stool affects children with obesity, we assessed its effect on several common Omega-3 fatty acids. Genetically proxied SM23-33 abundance in stool was significantly associated with the DHA [*OR*=1.059, (95%*CI*,1.012, 1.108), *P*=0.012], but not with ALA [*OR*=1.014, (95%*CI*, 0.855, 1.201), *P*=0.877], DPA [*OR*=1.057, (95%*CI*, 0.877, 1.275), *P*=0.559] and EPA [*OR*=1.043, (95%*CI*, 0.872, 1.248), *P*=0.646], and Genetically proxied ALA [*OR*=1.101, (95%*CI*, 0.974, 1.244), *P*=0.124], DPA [*OR*=1.022, (95%*CI*, 0.902, 1.159), *P*=0.729], EPA [*OR*=0.928, (95%*CI*, 0.850, 1.013, *P*=0.095] were not to be significantly associated with the children with obesity, but DHA was still significantly associated with children with obesity [*OR*=0.835, 95%*CI*, 0.735, 0.948, *P*=0.005], see Figure 4.

**Figure 4.**
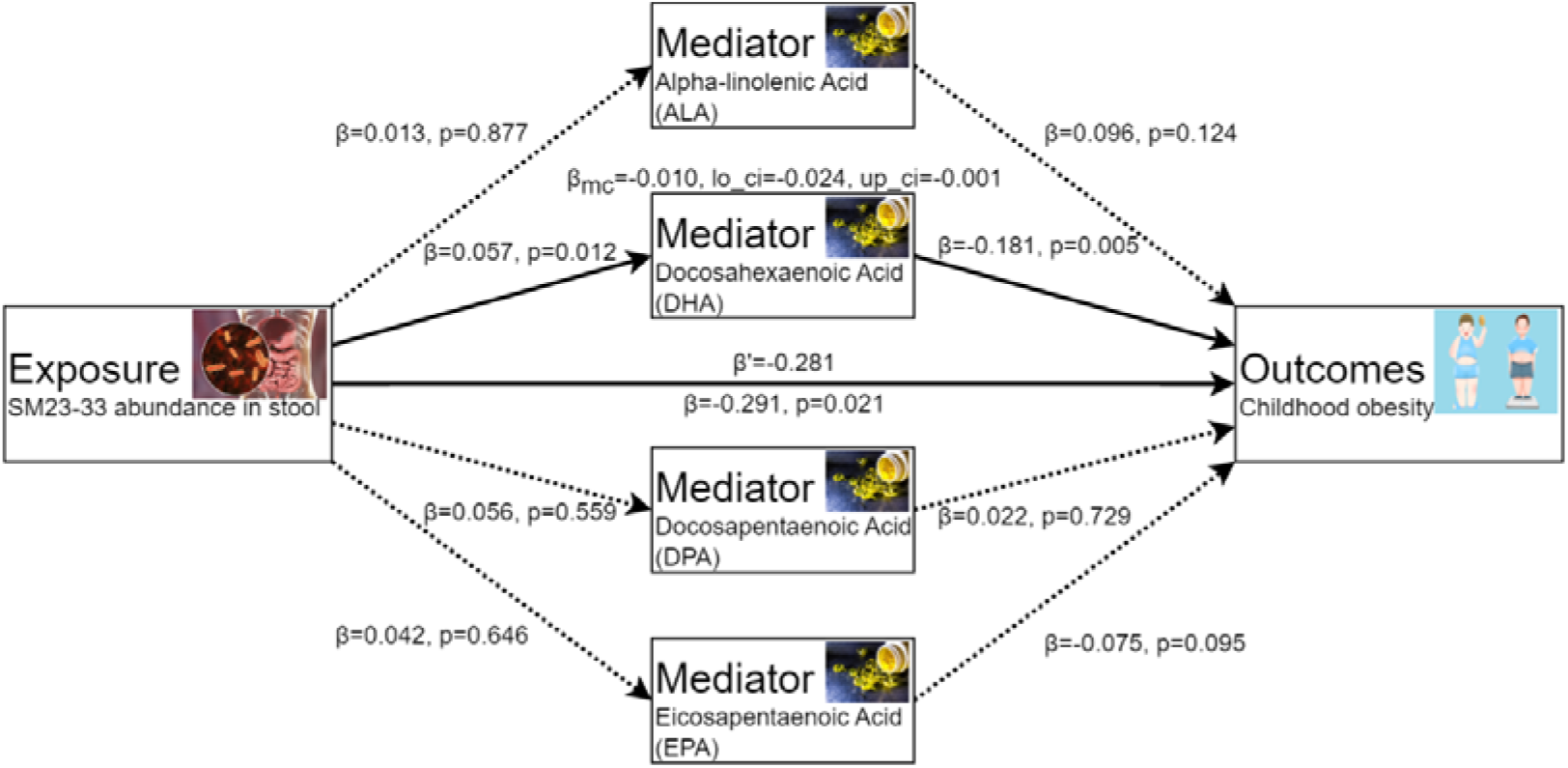
Schematic diagram of the mediating effect of Omega-3 fatty acids between SM23-33 abundance in Stool and SM23-33 abundance in Stool. The solid line represents that the causal relationship is established, and the dotted line represents that the causal relationship is not established.

The criteria for identifying mediators of the causal relationship between SM23-33 abundance in stool and children with obesity are as follows: a mediator must have a direct causal effect from SM23-33 abundance in stool, without any inverse relationships, and must also have a direct causal effect on children with obesity, and the relationship between SM23-33 abundance in stool and mediators, and the relationship between mediators and children with obesity should be in the same orientation. In line with the stated criteria, of the four analyzed omega-3 fatty acids (ALA, DHA, DPA, EPA), only DHA is a mediator between SM23-33 abundance in stool and children with obesity. More precisely, there is a positive causal association between SM23-33 abundance in stool and DHA [*OR*=1.059, (95%*CI*,1.012, 1.108), *P*=0.012], and DHA demonstrates negative causal links with children with obesity [*OR*=0.831, (95%*CI*, 0.719, 0.961), *P*=0.012]. We performed the mediation analysis to estimate the proportion of SM23-33 abundance in stool effect on children with obesity mediated through DHA. The mediation effect of DHA in the causal pathway from SM23-33 abundance in stool to children with obesity was - 0.010 (95%*CI*, -0.024, -0.001), accounting for 3.56% (95%*CI*, 3.43%, 3.69.0%) of the total effect. See Figure 4 and supplementary materials (Data S2, Table S5, S6, Figure S1, S2) for details.

### 3.5 Reverse MR analysis

First, children with obesity and Omega-3 fatty acids were used as exposures, and SM23-33 abundance in stool was used as the outcome. This direction of analysis aimed to investigate whether children with obesity and Omega-3 fatty acids have a causal effect on SM23-33 abundance in stool. Second, children with obesity was used as the exposure, and Omega-3 fatty acids were used as the outcome. This direction of analysis aimed to investigate whether children with obesity has a causal effect on Omega-3 fatty acids.

The results of the MR analysis found no evidence of a reverse causal link among SM23-33 abundance in stool, Omega-3 fatty acids, and children with obesity. In other words, the analysis did not suggest that children with obesity causes SM23-33 abundance in stool or Omega-3 fatty acids, or that Omega-3 fatty acids cause SM23-33 abundance in stool. The MR estimates and sensitivity analysis results are displayed in supplementary material (Data S2, Table S5, S7).

### 3.6 Sensitivity analysis

To address potential pleiotropy in our causal effect estimates, we performed multiple sensitivity analyses (see Table 2). Notably, Cochran’s Q-test and funnel plot analysis revealed no evidence of heterogeneity or asymmetry among the SNPs involved in the causal relationship. Furthermore, leave-one-out analysis validated the impact of each SNP on the overall causal estimates. Moreover, re-analysis of the MR study after excluding individual SNPs consistently yielded similar results, indicating that all SNPs contributed significantly to the establishment of the causal relationship. The relevant details are given in supplementary material (Data S2, Table S7, Figure S3).

**Table 2.**
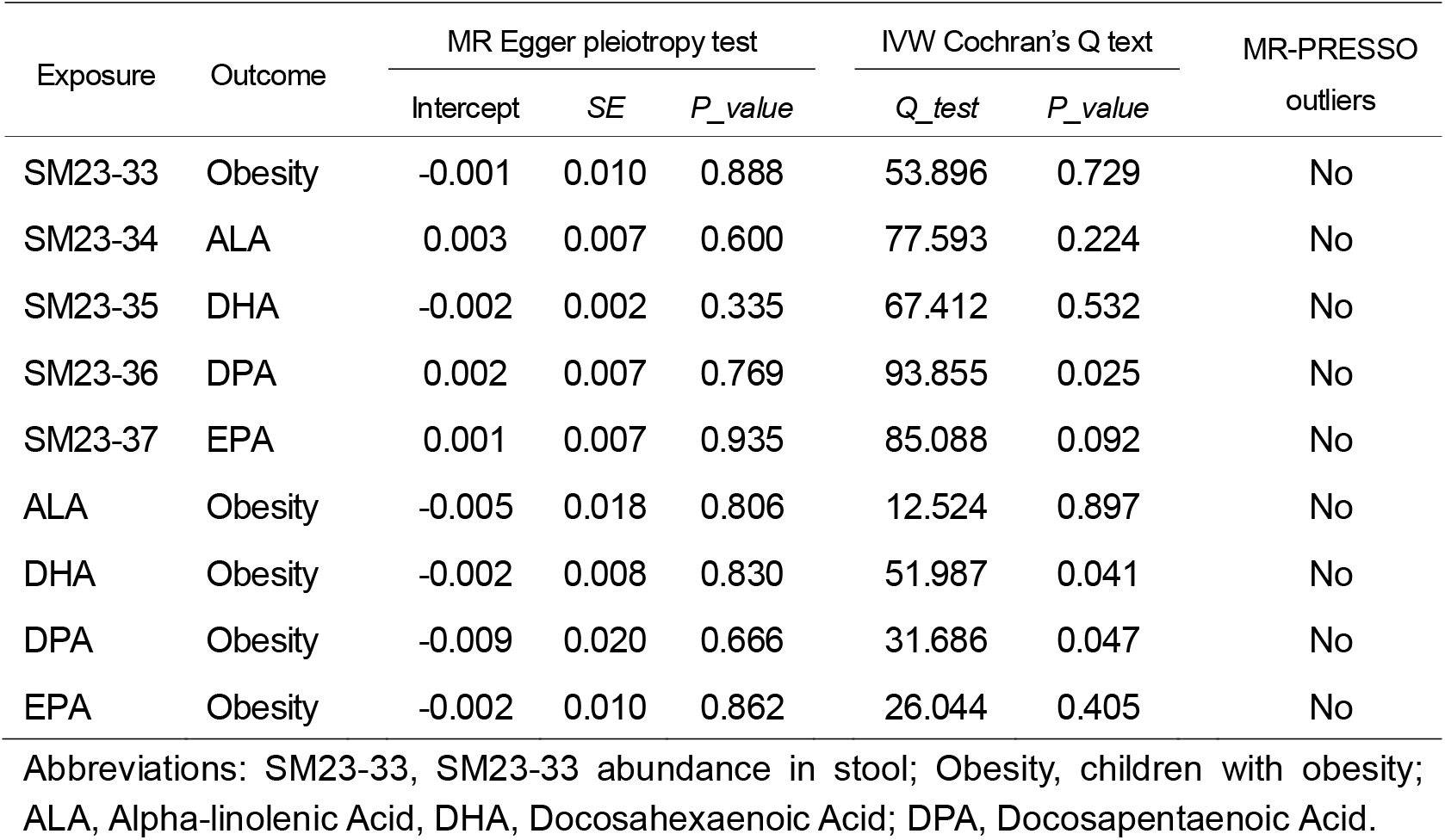
The results of the heterogeneity and horizontal pleiotropy.

## 4 DISCUSSION

In the present study, we investigated the causal association among SM23-33 abundance in stool, Omega-3 Fatty Acids, and children with obesity via large-scale genetic data and MR analysis. To our knowledge, the present study represents the first MR analysis to investigate the causal relationship among SM23-33 abundance in stool, Omega-3 Fatty Acids, and children with obesity. Through rigorous inclusion criteria and sensitivity analysis, we have identified potential causal links between SM23-33 abundance in stool and children with obesity, with one specific Omega-3 fatty acid serving as a mediator (DHA) in this pathway.

Studies have shown that the composition of intestinal flora in children with obesity is significantly different from that in children with healthy weight.^29^ In children with obesity, bacteria related to energy metabolism, such as Bacteroidetes and Clostridium are usually less abundant while bacteria linked to inflammation, such as Prevotella are relatively plentiful. ^30^ Due to the difficulty of separation, there is currently no research on the relationship between the abundance of SM23-33 in stool and children with obesity. This study found that the abundance of SM23-33 in stool may be an important intestinal bacteria for children with obesity, laying the foundation for further research about SM23-33 in the future.

Previous studies have shown that omega-3 fatty acids can reduce the body’s insulin resistance, affect the metabolism and storage of fat cells, reduce inflammatory responses and the risk of metabolic syndrome, and regulate appetite, lipid metabolism, and immune function,^31^ all of which may be related to inhibiting the occurrence of obesity in children. In this study, we selected four omega-3 fatty acids: ALA, DHA, DPA, and EPA. The results showed that only DHA helped to inhibit the occurrence of obesity in children (*OR*=0.747, 95%*CI*: 0.584-0.957; *P*=0.021) and played a weak mediating role in the role of SM23-33 abundance in stool in inhibiting children with obesity (3.56%, 95%CI: 3.43%, 3.69%). This shows that DHA may play a major role in the effect of omega-3 fatty acids on children with obesity. DHA has been proven to have many important health functions for the human body, such as promoting the development of the nervous system, benefiting cardiovascular health, regulating inflammation, improving the body’s resistance, ^32^ scavenging free radicals to reduce oxidative stress and cell damage,^33^ regulating blood lipids to prevent and treat chronic diseases,^34^ etc. If further research confirms the significant role of DHA in preventing and treating children with obesity, it may provide an effective approach to addressing this public health issue. Additionally, it could introduce novel and appropriate strategies for primary child healthcare facilities to use in combating children with obesity.

The possible mechanisms by which SM23-33 abundance leads to children with obesity include the following: (1) SM23-33 abundance in stool affects brain function and behavior through the gut-brain axis and may play a role in regulating appetite and energy metabolism, which also affects the occurrence of obesity in children.^35^ (2) Chronic low-grade intestinal inflammation can lead to the occurrence of insulin resistance and metabolic syndrome. SM23-33 abundance in stool inhibits the occurrence of chronic low-grade intestinal inflammation by protecting the intestinal barrier function and avoiding excessive activation of the immune system, thereby inhibiting the occurrence of obesity in children.^36^ (3) SM23-33 abundance in stool affects children with obesity by regulating energy metabolism. For example, some intestinal bacteria, such as Bacteroides, can break down complex carbohydrates and produce short-chain fatty acids, thereby promoting energy absorption and storage;^37^ Bifidobacteria and others can reduce fat accumulation by inhibiting energy absorption.^38^ (4) SM23-33 abundance in stool secretes various hormones that affect host metabolism and appetite regulation, thereby further inhibiting the occurrence of obesity in children. For example, some bacteria can produce leptin and leptin-like peptides, thereby regulating appetite and energy consumption.^39^ (5) SM23-33 abundance in stool affects host metabolism and energy regulation by producing metabolites produced by certain microorganisms, such as short-chain fatty acids and bile acids, further affecting the development of obesity in children.^40^

The significance of SM23-33 abundance in stool in the prevention and treatment of children with obesity may include: (1) Probiotics are live microorganisms that provide health benefits to the host when ingested in sufficient amounts. Prebiotics, such as inulin and fructan, are indigestible substances that promote the growth of beneficial bacteria in the intestine.^41^ The production of probiotics and prebiotics related to SM23-33 further regulates the composition of intestinal flora and improves intestinal health, which may help prevent children with obesity. (2) Fecal flora transplantation is the transfer of fecal flora from healthy donors to the intestines of recipients to restore healthy flora. Fecal flora transplantation has made some progress in obesity,^42^ and SM23-33 may become a new effective therapeutic target。

The link between gut microbiota and children with obesity is complex and requires further study. Future research should focus on clarifying the exact mechanism by which gut microbiota causes obesity in children and developing truly effective interventions.

The advantages of our study are that we used a large GWAS database and adopted a robust MR analysis method to conduct the study. In order to ensure the rigor of the study, we used a variety of research methods, such as genetic correlation regression analysis (LDSC) and reverse MR analysis of all paths. However, this study also has some disadvantages. First, although we applied MR-PRESSO and MR-Egger methods to evaluate the impact of heterogeneity and pleiotropy on the risk estimation of MR analysis, and we used the LDlink website to remove SNPs that might be confounding factors, since the study of SNPs is always ongoing, we cannot completely rule out possible horizontal pleiotropy, which may lead to biased causal effect estimates. Second, although the SNPs in our study were extracted from the existing large GWAS, there may be certain heterogeneity between different GWAS databases, which will also affect the accuracy of the study. Third, the genetic variation in our study was mainly from individuals of European ancestry, but there is still the possibility of interference from a small number of participants of other ethnicities, which may cause a slight bias and affect the generalizability of the results, and also limit the applicability of the results to other ethnic groups. Fourth, the intestinal flora may be affected by factors such as demographics, diet, or medication, most of which have heterogeneity, inter-individual variability, and low heritability, which may reduce the statistical power and robustness of the results.

In conclusion, the present study is the first to comprehensively assess the causal relationships between SM23-33 abundance in stool, Omega-3 fatty acids, and children with obesity, emphasizing DHA of Omega-3 fatty acids in this disease. The results offer important insights into the etiology of children with obesity and present novel insights for therapeutic and preventive strategies. Furthermore, we briefly elucidated the possible underlying mechanisms between SM23-33 abundance in stool and children with obesity. These results provide new insights into gut microbiota treatment of children with obesity.

## Data Availability

The data used in this study are available from the corresponding author upon reasonable request. The analysis utilized publicly available datasets, and we extend our highest respect to the data providers. Detailed information on all original contributions can be found in the article and accompanying Supplementary Information. For further inquiries, please get in touch with the corresponding author.

https://www.ebi.ac.uk/gwas

## ACKNOWLEDGEMENTS

This study was independently conducted by the author, who confirms that no other individuals meeting the criteria were omitted. The author has reviewed and approved the final version of the manuscript submitted for publication.

